# Systematic Review and Meta-Analysis of Sex-Specific COVID-19 Clinical Outcomes

**DOI:** 10.1101/2020.05.11.20098673

**Authors:** Thushara Galbadage, Brent M. Peterson, Joseph Awada, Alison S. Buck, Danny A. Ramirez, Jason Wilson, Richard S. Gunasekera

## Abstract

To successfully mitigate the extraordinary devastation caused by the Coronavirus disease 2019 (COVID-19) pandemic, it is crucial to identify important risk factors for this disease. One such neglected health determinant is the sex of the patient. This is an essential clinical characteristic, as it can factor into a patient’s clinical management and preventative measures. Some clinical studies have shown disparities in the proportion between males and females that have more severe clinical outcomes or, subsequently, die from this disease. However, this association has not been unequivocally established. Thus, the purpose of this investigation was to examine the association between male sex and COVID-19 severity. We systematically reviewed the literature, identified non-randomized studies that matched predetermined selection criteria, and performed a meta-analysis to evaluate the proportion of males among four disease severity categories. Appropriate assessment strategies were implemented to assess and minimize potential biases. The results of this meta-analysis indicated that males constituted a significantly higher proportion of those who had adverse clinical outcomes and died from COVID-19. As the coronavirus spread from the East to the West, male sex remained a consistent risk factor. Our results support the establishment of the male sex as an important risk factor for this disease. Early identification and appropriate medical care for males with lab-confirmed COVID-19 may substantially change the course of clinical prognosis, resulting in greater numbers of lives saved.

## 1 Introduction

Males and females have distinct biological, immunological, and endocrine differences that result in different disease processes and outcomes. Sex-specific differential gene expression and molecular-level variation have been reported to influence blood pressure, cardiovascular health, and kidney function (Convertino, 1998; Reckelhoff, 2001; Fischer et al., 2002; Kang and Miller, 2002; Sandberg and Ji, 2003; Hilliard et al., 2013). Females, in general, have a heightened capability to activate a greater and more robust immune response, offering protection against many infectious disease processes, but may predispose them to an array of autoimmune diseases (Fairweather and Rose, 2004; Pennell et al., 2012; Rubtsova et al., 2015; Klein and Flanagan, 2016; vom Steeg and Klein, 2016; Jaillon et al., 2019). Males and females also express immunological dimorphisms. Females have two X chromosomes in comparison to the XY in males. The random transcriptional inactivation of X chromosomes in females may also help offset certain mutation-related dysregulation of the immune system (Taneja, 2018). Differences in endocrine system regulation in females compared to males significantly affect disease processes including respiratory, cardiovascular, and renal disease (Hilliard et al., 2013; Blenck et al., 2016; Channappanavar et al., 2017; Palmisano et al., 2018; Vermillion et al., 2018; Wensveen et al., 2019). As nations across the world navigate their way through the Coronavirus disease 2019 (COVID-19) pandemic, clinical, research, and public health experts have observed that this disease does not affect all individuals alike.

Since the beginning of 2020, the world’s healthcare professionals have tirelessly attempted to mitigate the impact of the COVID-19 pandemic. With over 3.5 million confirmed cases and 243,000 deaths worldwide as of May 5th, 2020, a post-COVID-19 pandemic era is not within the near foreseeable future (WHO, 2020). The United States, one of the epicenters for the disease, has documented over 1.1 million confirmed cases and 68,000 deaths related to COVID-19 (CDC, 2020; WHO, 2020). Many recent studies have highlighted certain risk factors that cause specific populations to be disproportionately susceptible to the severe acute respiratory syndrome coronavirus 2 (SARS-CoV-2) infection. Currently known risk factors for severe clinical outcomes of COVID-19 include: advanced age (65 years and older), chronic lung diseases, immunocompromised status, and other comorbidities such as hypertension, diabetes, and/or cardiovascular disease (Emami et al., 2020; Grasselli et al., 2020; Remuzzi and Remuzzi, 2020; Shahid et al., 2020; Zheng et al., 2020).

Observations in COVID-19 patient data involving clinical characteristics highlight specific disparities in males and females. A recent case-series study looking at COVID-19 and SARS patients showed that while males and females had the similar disease prevalence, males with COVID-19 were at higher risk for worse clinical outcomes and death (Jin et al., 2020). In this study, as the patient age and the documented comorbidities (i.e., cardiovascular diseases, diabetes, chronic lung diseases, or hypertension) increased, the risk of severity and mortality in both COVID-19 and SARS patients increased. However, the mortality rate in males was 2.4 times that of their age-matched female counterparts (70.3% and 29.7%, respectively).

Furthermore, a nationwide COVID-19 surveillance study conducted in Italy indicated that male mortality rates related to COVID-19 were disproportionately higher than that of female patients with a ratio as much as 4 to 1 (Remuzzi and Remuzzi, 2020). Other systematic reviews performed to characterize clinical features or risk factors for COVID-19, have also identified the sex-specific disparities in disease severity and mortality (Li et al., 2020a; Zheng et al., 2020). However, the clinical importance of male sex as a risk factor for COVID-19 has mainly been overlooked or explained as a potential confounder to other environmental factors such as smoking or tobacco product usage (Cai, 2020). While various studies have made observations of the sex-specific disparities of COVID-19, this specific relationship has not been adequately established. The sex-specific disease severity is an important clinical consideration as it affects all patient populations. Recognition of male sex as a risk factor for COVID-19 will impact both preventative measures and clinical patient management protocols.

The goal of this systematic review and meta-analysis is to identify whether males are more susceptible to COVID-19, severe forms of the disease, or mortality related to COVID-19. To address this question, we systematically reviewed the literature using the Preferred Reporting Items for Systematic Reviews and Meta-Analyses (PRISMA) guidelines. We performed a meta-analysis of the selected study populations comparing male and female COVID-19 patients. This review incorporated three online databases and research studies published between December 15th, 2019, and April 16th, 2020. We characterized the influence of sex as a risk factor for COVID-19 measuring the following clinical outcomes: all lab-confirmed cases, severe cases, critically ill cases, and mortality.

## 2. Methods

### 2.1 Literature Search and Research Study Selection

We performed a comprehensive systematic research literature search of three online databases, PubMed (LitCOVID), Embase (OVID), and Web of Science (WoS), from December 15th, 2019, to April 16th, 2020. We identified all research articles related to COVID-19 that contained any sex-specific patient or clinical characterizations. The search terms and keywords used to identify research studies for the meta-analysis were: COVID-19, male, female, men, women, sex, and gender. We reviewed references of review, perspectives, systematic reviews, and meta-analysis articles of the include articles to ensure comprehensiveness of our search. All our search results were evaluated using the PRISMA statement. We reviewed the abstracts and tables of each of the articles to identify the presence of sex-specific (male and female) COVID-19 case numbers.

### 2.2 Eligibility criteria

The inclusion criteria for research article selection was as stated below. Study population: patients with lab-confirmed COVID-19 diagnosis. Study design: case series or cross-sectional study that did not exclude any lab-confirmed COVID-19 patients. Outcomes measure: at least one outcome reported with male to female ratio among lab-confirmed clinical cases, severe cases, critical cases, and mortality. Research study: only peer-reviewed research publications were included. Commentary articles, perspectives, review articles, and surveillance reports were excluded. The following case definitions were used in this study. All cases were lab-confirmed COVID-19 patients. Severe cases were defined as having at least one of the following clinical findings: (a) breathing rate ≥30/min, (b) oxygen saturation (SpO2) ≤ 93% at rest, or (c) ratio of the partial pressure of arterial oxygen (PaO2) to the fraction of inspired oxygen (FiO2) ≤300 mmHg. The severe case definition followed the American Thoracic Society guidelines for community-acquired pneumonia (Metlay et al., 2019). Critical cases were defined as: (a) received mechanical ventilation; (b) clinically diagnosed with shock symptoms, (c) received care in the intensive care unit (ICU) or (d) transfer to a higher level of medical care.

### 2.3 Data Extraction and Quality Assessment

All articles identified through the keyword search from the online databases were organized into an Excel^®^ spreadsheet. Following the removal of duplicates, articles were subjected to evaluation, and five investigators did data extraction. Research studies were screened using the abstract and any tabulated clinical characteristics of COVID-19 patients. Directly after that, research articles were again screened to identify any discrepancies by an independent investigator. The screened articles were assessed against the study selection criteria by two independent investigators, and any differences in selected articles were revisited, and a definitive determination was made.

### 2.4 Selected Study Bias Risk Assessment

A bias risk assessment was conducted on studies included in the meta-analysis utilizing the methodological index for non-randomized studies (Minors) criteria at the study level (Slim et al., 2003). Each of the selected articles was scored with 0 (not reported), 1 (reported but inadequate), or 2 (reported and adequate). The highest score possible was 16 for non-comparative studies according to Minors guidelines.

### 2.5 Statistical Analysis of Selected Data Sets

Statistical analysis was conducted using *R* (R_Core_Team, 2019) with the meta-analysis packages *meta* (Balduzzi et al., 2019) and *dmetar* (Harrer et al., 2019). The principal summary measures of the meta-analysis were proportions of males in four different COVID-19 categories. The four groups were: (a) all confirmed COVID-19 cases, (b) severe cases of COVID-19 as defined in Section 2.3, (c) critically ill cases of COVID-19 as defined in Section 2.3, and (d) deaths associated with COVID-19. Agresti-Coull confidence intervals were used for individual studies. Studies were combined using the inverse variance method on the raw proportions with the DerSimonian-Laird estimator for the between-groups variance estimator (*τ*^2^) and the Jackson method for combined confidence intervals. Transformations of raw proportions were calculated for the combined estimates (*log, logit, arcsin*, and *Freeman-Tukey double arcsin*), but the results were so similar they are not shown. The proportion of variation in treatment effects was estimated with *I^2^*. To assess bias across studies, funnel plots were constructed for each of the four different categories, and Egger’s bias test conducted. In order to determine if there were region-specific differences among populations in Asian and Western countries, we sub-divided the COVID-19 critically ill patient populations into these two regions and analyzed them.

### 2.6 Clinical Outcomes Median Age Calculation

To combine the ages, in 20 of the articles, the median age of patients was given, along with sometimes interquartile range, sometimes min and max. In 10 of the articles, mean and standard deviation (SD) were presented. In one article (Easom et al., 2020), the mean age was given without SD. We used linear regression on the other 10 (mean, SD) pairs to estimate the SD to be 14.5 years. To combine the ages, we chose to convert means to medians because there would be fewer unknown statistics to estimate, and typical disease distributions are skewed. To convert, we fit a negative binomial distribution to the mean and SD using the method of moments. With the complete list of medians, we used *R*’s *metamedian* (McGrath et al., 2020) package to obtain summarized confidence intervals for each of the four categories.

## 3. Results

### 3.1 Research Study Selection and Quality Assessment

We identified 786 research articles that matched our search terms. After the duplicated were removed, 414 unique research articles were screened. Following the screening process, 353 articles with incomplete data were excluded. We then identified 61 research articles with sex-specific case numbers and reviewed full-length articles to assess their eligibility for our study according to the selection criteria. Thirty articles did not fit the selection criteria and were excluded from the meta-analysis. Reasons for exclusion were: not a primary research study (a surveillance report or perspective), did not include consecutive patients or did not meet with the case or severity definitions. The 31 research articles eligible for this meta-analysis were used for qualitative synthesis and quantitative analysis (Figure 1). The 31 eligible articles were subjected to a bias assessment using the Minors criteria at the study level (Slim et al., 2003). All 31 selected articles scored between 12 and 14 points, with 16 being the highest for non-randomized controlled studies (Table 1). The relatively high scores indicated that we were likely not introducing any significant systemic biases.

**Figure 1.**
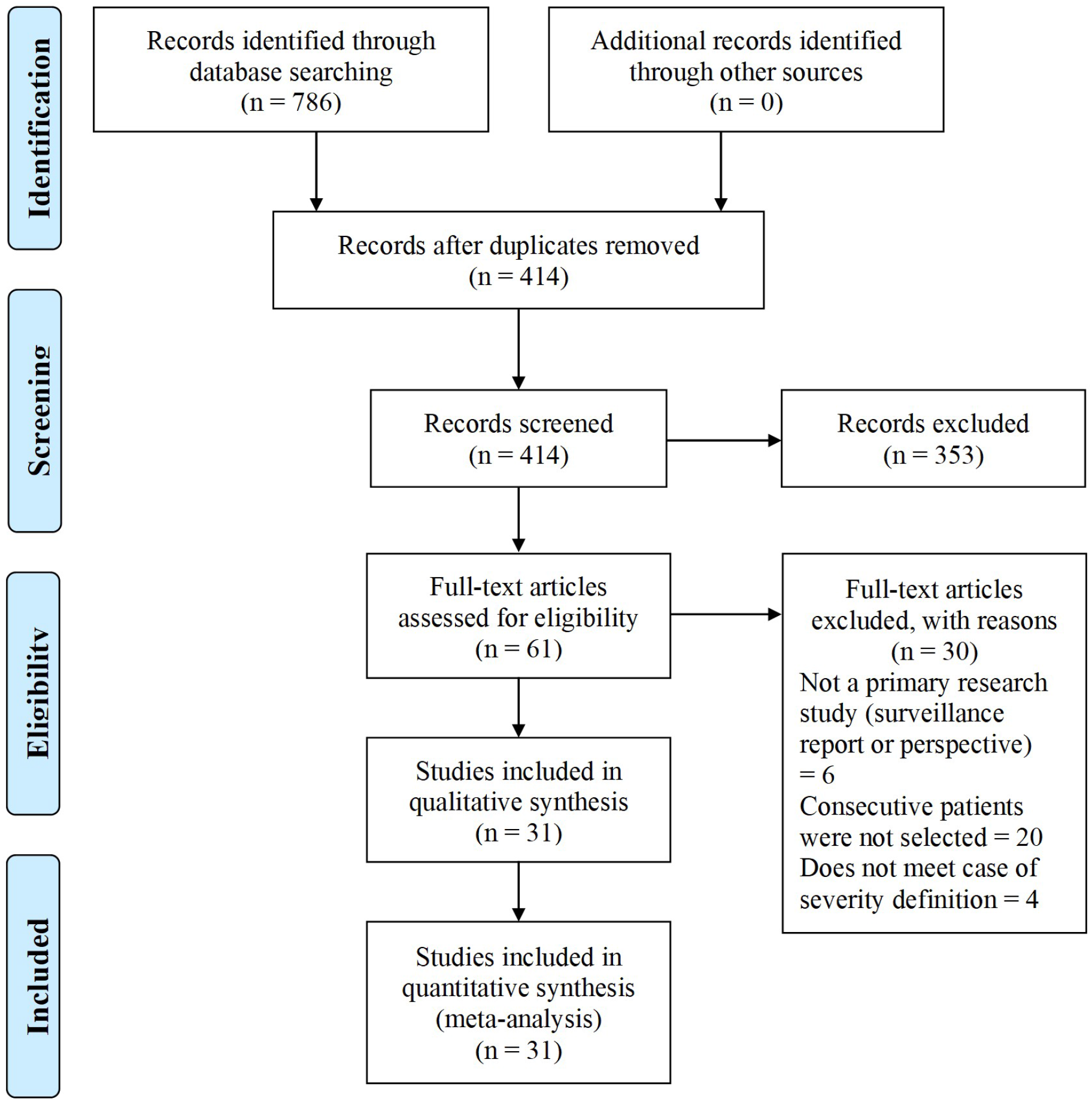
COVID-19 sex-specific disease severity flow diagram of the inclusion criteria of studies eligible for meta-analysis. Flow diagram template adopted from the PRISMA approach to meta-analysis (Moher et al., 2009).

**Table 1.** Bias risk assessment on the studies included in the meta-analysis using the methodological index for non-randomized studies (Minors) criteria (Slim et al., 2003).

| Study Reference | Study Population | (1) | (2) | (3) | (4) | (5) | (6) | (7) | (8) | Score |
| --- | --- | --- | --- | --- | --- | --- | --- | --- | --- | --- |
| Barrasa et al., 2020 | 48 | 2 | 2 | 2 | 2 | 2 | 2 | 2 | 0 | 14 |
| Bhatraju et al., 2020 | 24 | 2 | 2 | 2 | 2 | 2 | 1 | 1 | 0 | 12 |
| Cao et al., 2020 | 102 | 2 | 2 | 2 | 2 | 2 | 1 | 1 | 0 | 12 |
| Chen et al., 2020a | 249 | 2 | 2 | 2 | 2 | 2 | 1 | 1 | 0 | 12 |
| Chen et al., 2020b | 99 | 2 | 2 | 2 | 2 | 2 | 2 | 2 | 0 | 14 |
| Chen et al., 2020c | 203 | 2 | 2 | 2 | 2 | 2 | 2 | 2 | 0 | 14 |
| Chen et al., 2020d | 113 | 2 | 2 | 2 | 2 | 2 | 1 | 2 | 0 | 13 |
| Chu et al., 2020 | 54 | 2 | 2 | 2 | 2 | 2 | 1 | 1 | 0 | 12 |
| Du et al., 2020a | 179 | 2 | 2 | 2 | 2 | 2 | 2 | 1 | 0 | 13 |
| Du et al., 2020b | 109 | 2 | 2 | 2 | 2 | 2 | 2 | 2 | 0 | 14 |
| Du et al., 2020c | 85 | 2 | 2 | 2 | 2 | 2 | 2 | 2 | 0 | 14 |
| Easom et al., 2020 | 68 | 2 | 2 | 2 | 2 | 2 | 2 | 2 | 0 | 14 |
| Grasselli et al., 2020 | 1591 | 2 | 2 | 2 | 2 | 2 | 2 | 2 | 0 | 14 |
| Guan et al., 2020 | 1096 | 2 | 2 | 2 | 2 | 2 | 1 | 1 | 0 | 12 |
| Huang et al., 2020 | 41 | 2 | 2 | 2 | 2 | 2 | 1 | 1 | 0 | 12 |
| Korea, 2020 | 54 | 2 | 2 | 2 | 2 | 2 | 2 | 2 | 0 | 14 |
| Li et al., 2020b | 548 | 2 | 2 | 2 | 2 | 2 | 1 | 2 | 0 | 13 |
| Liu et al., 2020 | 137 | 2 | 2 | 2 | 2 | 2 | 2 | 2 | 0 | 14 |
| Mao et al., 2020 | 214 | 2 | 2 | 2 | 2 | 2 | 1 | 1 | 0 | 12 |
| Qin et al., 2020 | 452 | 2 | 2 | 2 | 2 | 2 | 1 | 1 | 0 | 12 |
| Simonnet et al., 2020 | 85 | 2 | 2 | 2 | 2 | 2 | 1 | 1 | 0 | 12 |
| Wan et al., 2020 | 135 | 2 | 2 | 2 | 2 | 2 | 1 | 1 | 0 | 12 |
| Wang et al., 2020a | 138 | 2 | 2 | 2 | 2 | 2 | 1 | 1 | 0 | 12 |
| Wang et al., 2020b | 125 | 2 | 2 | 2 | 2 | 2 | 2 | 2 | 0 | 12 |
| Wang et al., 2020c | 1012 | 2 | 2 | 2 | 2 | 2 | 1 | 2 | 0 | 13 |
| Wu et al., 2020 | 80 | 2 | 2 | 2 | 2 | 2 | 2 | 2 | 0 | 14 |
| Xie et al., 2020 | 79 | 2 | 2 | 2 | 2 | 2 | 1 | 1 | 0 | 12 |
| Xu et al., 2020 | 90 | 2 | 2 | 2 | 2 | 2 | 2 | 2 | 0 | 14 |
| Yang et al., 2020 | 149 | 2 | 2 | 2 | 2 | 2 | 2 | 2 | 0 | 14 |
| Young et al., 2020 | 18 | 2 | 2 | 2 | 2 | 2 | 2 | 2 | 0 | 14 |
| Zhang et al., 2020 | 140 | 2 | 2 | 2 | 2 | 2 | 1 | 1 | 0 | 12 |

### 3.2 Study Population Demographics

Within our selected studies, 7556 lab-confirmed COVID-19 cases were identified. Of these 31 studies, 24 were from various cities in China and included a sample of 5629 lab-confirmed cases. Two studies were from South Korea and Singapore, which included a sample of 72 lab-confirmed cases. The other five studies were from Europe and North America, having a sample of 1855 lab-confirmed cases (Figure 2 and Table 2). Most of the early studies came from China with study periods from December 11th, 2019, to February 24th, 2020. Most of the later studies came from other countries with study periods from January 23rd to April 5th, 2020 (Figure 3). These patterns reflect the movement of epicenters for COVID-19 from the East to the West.

**Figure 2.**
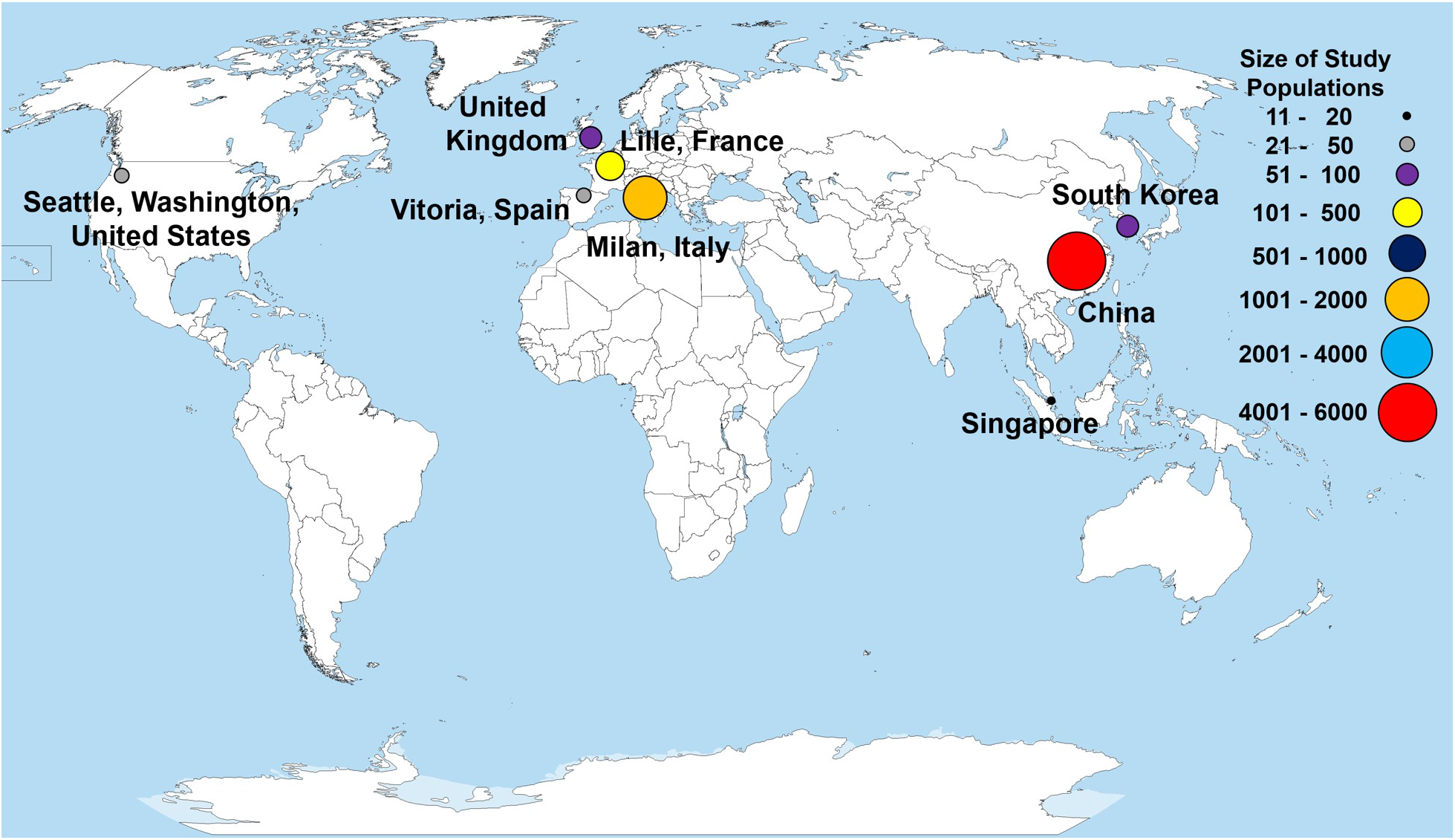
Countries and locations for the selected studies used in the meta-analysis. Total patient populations in each of the study locations are illustrated with a colored circle and correspond to the size of study populations. Each point represents a research study, except for China, which represents the total patient population from 24 different studies. The world map was obtained from Wikimedia Commons, the free media repository licensed under the Creative Commons Attribution-Share Alike 3.0 Unported license.

**Table 2.** Demographics of all studies included in the meta-analysis with sex-specific disease severity.

| Study Reference | Country<br>(City or Province) | Study<br>Population | Age <sup>1</sup><br>(*mean* or<br>*median*) | All<br>cases <sup>2</sup><br>(male %) | Severe<br>cases <sup>3</sup><br>(male %) | Critical<br>cases <sup>4</sup><br>(male %) | Mortality<br>(male %) |
| --- | --- | --- | --- | --- | --- | --- | --- |
| Barrasa et al., 2020 | Spain (Vitoria) | 48 | 63.2 | - | - | 56.3 | - |
| Bhatraju et al., 2020 | United States<br>(Seattle) | 24 | 64.0 | - | - | 62.5 | - |
| Cao et al., 2020 | China (Wuhan) | 102 | 54.0 | 52.0 | - | - | 76.5 |
| Chen et al., 2020a | China (Shanghai) | 249 | 51.0 | 50.6 | - | 86.4 | - |
| Chen et al., 2020b | China (Wuhan) | 99 | 55.5 | 67.7 | - | - | - |
| Chen et al., 2020c | China (Wuhan) | 203 | 54.0 | 53.2 | - | - | - |
| Chen et al., 2020d | China (Wuhan) | 113 | 68.0 | - | - | - | 73.5 |
| Chu et al., 2020 | China (Wuhan) | 54 | 39.0 | 66.7 | 69.8 | - | - |
| Du et al., 2020a | China (Wuhan) | 179 | 57.6 | 54.2 | - | - | 47.6 |
| Du et al., 2020b | China (Wuhan) | 109 | 70.7 | - | - | - | 67.9 |
| Du et al., 2020c | China (Wuhan) | 85 | 65.8 | - | - | - | 72.9 |
| Easom et al., 2020 | United Kingdom | 68 | 42.5 | 47.1 | - | - | - |
| Grasselli et al., 2020 | Italy (Milan) | 1591 | 63.0 | - | - | 82.0 | - |
| Guan et al., 2020 | China | 1096 | 47.0 | 58.1 | 57.8 | 67.2 | - |
| Huang et al., 2020 | China (Wuhan) | 41 | 49.0 | 73.2 | - | 84.6 | - |
| Korea, 2020 | South Korea | 54 | 75.5 | - | - | - | 61.1 |
| Li et al., 2020b | China (Wuhan) | 548 | 60.0 | 50.9 | 56.9 | - | - |
| Liu et al., 2020 | China (Wuhan) | 137 | 57.0 | 44.5 | - | - | - |
| Mao et al., 2020 | China (Wuhan) | 214 | 52.7 | 40.7 | 50.0 | - | - |
| Qin et al., 2020 | China (Wuhan) | 452 | 58.0 | 52.0 | 54.2 | - | - |
| Simonnet et al., 2020 | France (Lille) | 124 | 60 | - | - | 72.6 | - |
| Wan et al., 2020 | China (Chongqing) | 135 | 47.0 | 53.3 | 52.5 | - | - |
| Wang et al., 2020a | China (Wuhan) | 138 | 56.0 | 54.3 | - | 61.1 | - |
| Wang et al., 2020b | China (Fuyang) | 125 | 38.8 | 56.8 | - | - | - |

COVID-19 Sex-Specific Clinical Outcomes
| Study Reference | Country<br>(City or Province) | Study<br>Population | Age <sup>1</sup><br>(*mean* or<br>median) | All<br>cases <sup>2</sup><br>(male %) | Severe<br>cases <sup>3</sup><br>(male %) | Critical<br>cases <sup>4</sup><br>(male %) | Mortality<br>(male %) |
| --- | --- | --- | --- | --- | --- | --- | --- |
| Wang et al., 2020c | China (Wuhan) | 1012 | 50.0 | 51.8 | - | 62.0 | - |
| Wu et al., 2020 | China (Jiangsu) | 80 | *46.1* | 48.8 | - | - | - |
| Xie et al., 2020 | China (Wuhan) | 79 | 60.0 | 55.7 | 64.3 | - | - |
| Xu et al., 2020 | China (Wuhan) | 90 | 50.0 | 43.3 | - | - | - |
| Yang et al., 2020 | China (Wenzhou) | 149 | *45.1* | 54.4 | - | - | - |
| Young et al., 2020 | Singapore | 18 | 47.0 | 50.0 | - | - | - |
| Zhang et al., 2020 | China (Wuhan) | 140 | 57.0 | 50.7 | 56.9 | - | - |

<sup>1</sup> Mean or median age of the study population for each research study. In the event a study had a severity or mortality sub-population, age is listed for only the total study population. Mean ages are indicated in *italics*.
<sup>2</sup> All consecutive patients with lab confirmed case of COVID-19 within the study period.
<sup>3</sup> Severe case defined as having at least one of the following clinical findings: (a) breathing rate $\geq 30/\text{min}$ , (b) pulse oximeter oxygen saturation ( $\text{SpO}_2$ ) $\leq 93\%$ at rest, or (c) ratio of partial pressure of arterial oxygen ( $\text{PaO}_2$ ) to fraction of inspired oxygen ( $\text{FiO}_2$ ) $\leq 300$ mmHg
<sup>4</sup> Critical case defined as: (a) received mechanical ventilation; (b) clinically diagnosed with shock, (c) received care in the intensive care unit (ICU) or (d) transferred to a higher level of medical care

**Figure 3.**
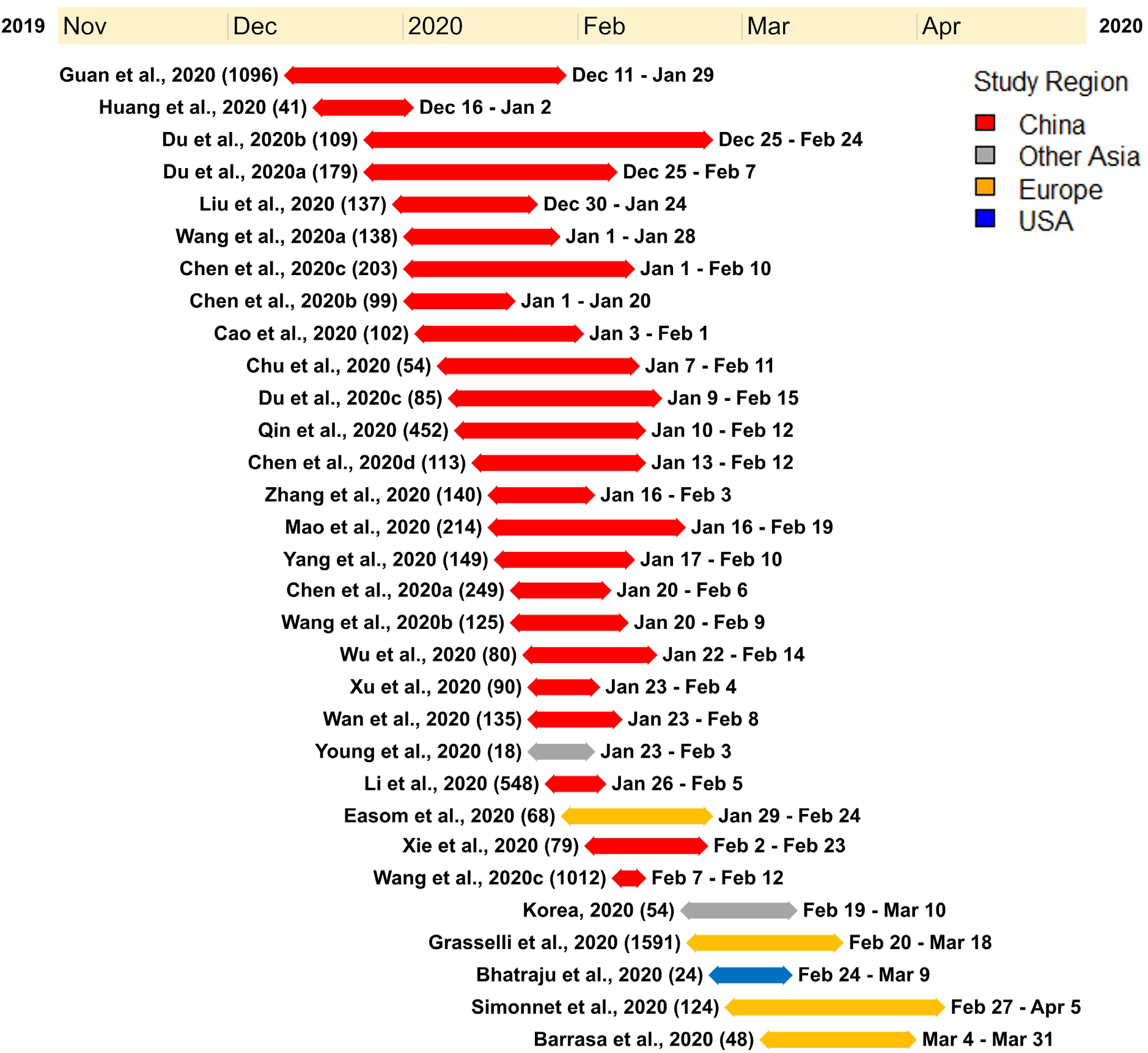
A timeline illustrating the study period of each of the research studies used for the meta-analysis. Each research study used for the meta-analysis is represented by the study name, (study sample), duration of the study with a line corresponding to the length of the study, and the start and end date of the study. The studies were ordered according to the start date of each study.

### 3.3 Meta-Analysis and Bias Assessment

The principal quantitative results are contained in the forest plots shown on the left side of Figures 4 and 5. The individual confidence intervals are shown, by study, with the combined proportion for each group and confidence interval at the bottom. A random-effects model was used for the combined proportion to check for heterogeneity (*τ*^2^= between-group variation and *I*^2^= proportion of total variation in the estimates of treatment effects due to heterogeneity). The heterogeneity statistics (*τ*^2^ and *I^2^*) are shown at the bottom left of the forest plots.

**Figure 4.**
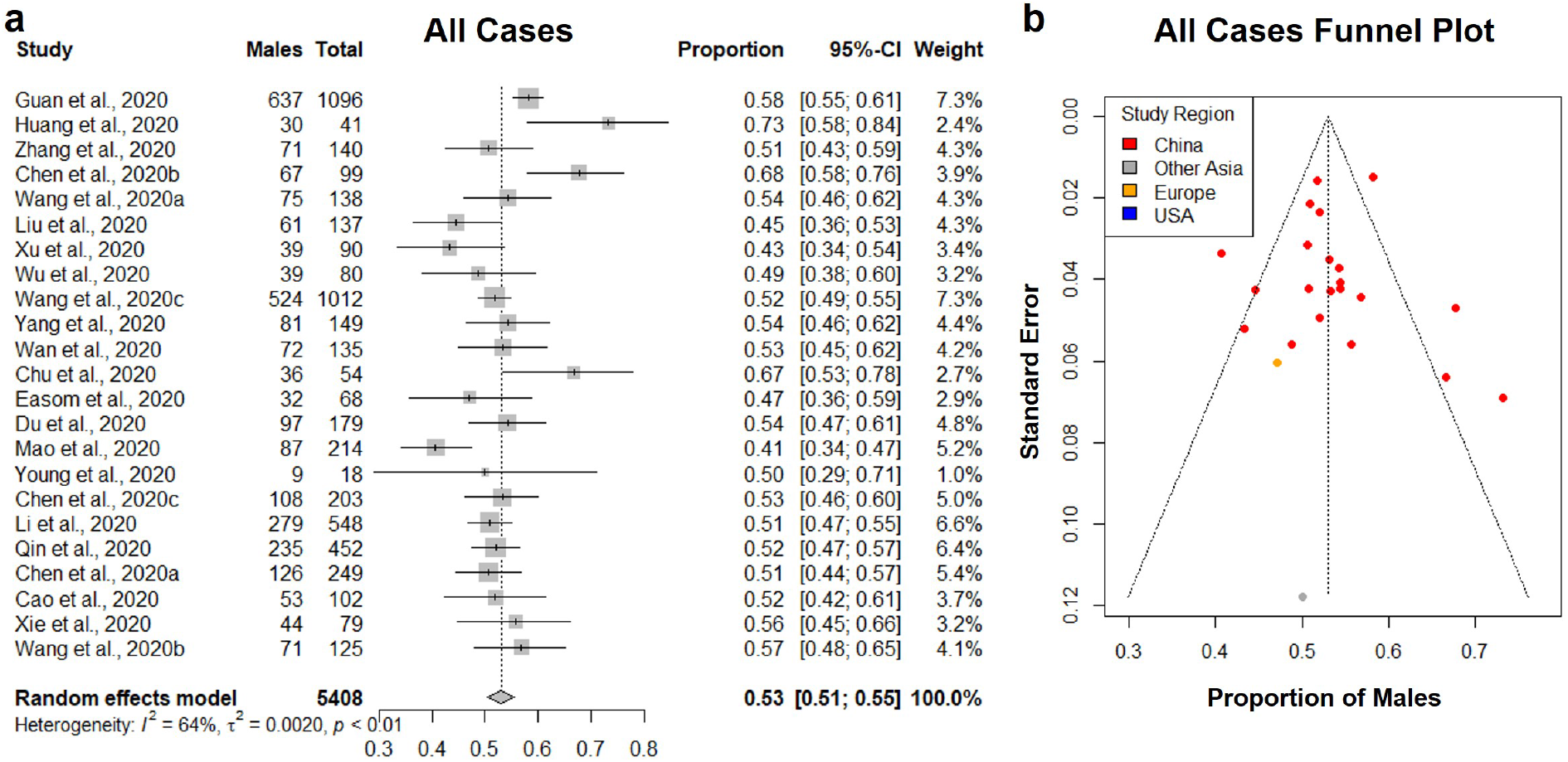
Proportion of males in all lab-confirmed COVID-19 cases. (a) Forest plot of sexdistribution in all lab-confirmed COVID-19 cases in each of the studies. Proportions of males and the 95% confidence intervals are indicated. (b) Funnel plot with 95% confidence region of sexdistribution in all lab-confirmed COVID-19 cases in each of the studies.

**Figure 5.**
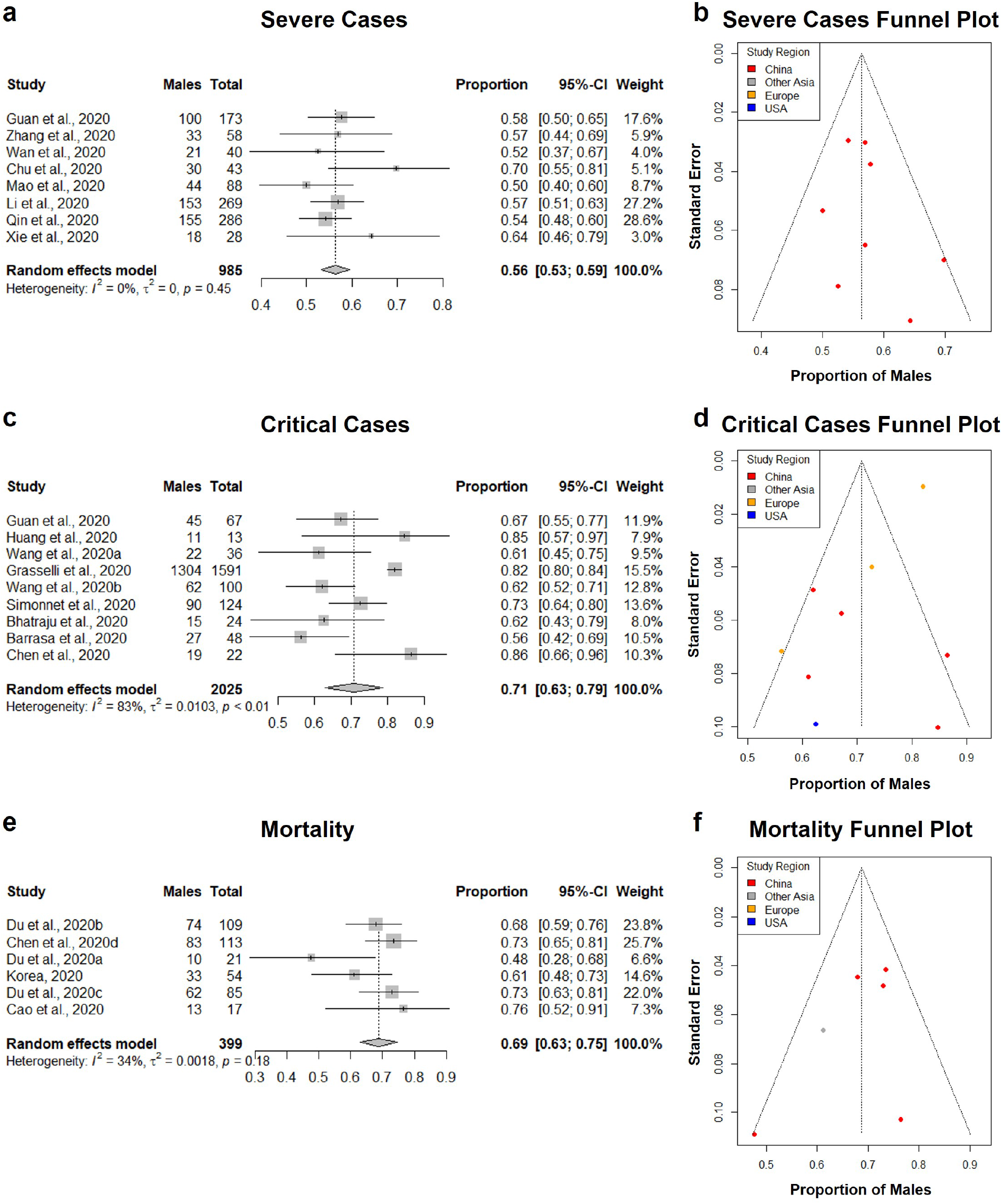
Proportion of males in COVID-19 severe cases, critical cases, and mortalities. (a, c, e) Forest plot of sex-distribution in COVID-19 cases in each of the studies. Proportions of males and the 95% confidence intervals are indicated. (a) Severe cases defined as having at least one of the following clinical findings: breathing rate ≥30/min, pulse oximeter oxygen saturation (SpO2) ≤93% at rest, or ration of the partial pressure of arterial oxygen (PaO2) to the fraction of inspired oxygen (FiO2) ≤300 mmHg. (c) Critical case defined as: received mechanical ventilation, clinically diagnosed with shock, received care in the intensive care unit (ICU), or transferred to a higher level of medical care. (e) Mortality defined as all deaths in COVID-19 patients that occurred during the study period. (b, d, f) Funnel plot with 95% confidence region of sex-distribution in COVID-19 severe cases, critical cases, and mortality in each of the studies.

### 3.4 Sex-Specific COVID-19 Case Distribution

A total of 23 studies with 5408 lab-confirmed COVID-19 cases were analyzed (Table 3). Our results from the randomized effects model meta-analysis showed that in the sex-distribution of all COVID-19 cases, males accounted for 53% (95% CI [0.51, 0.55]) (Figure 4a). Female patients made up 47% of all COVID-19 cases. There is moderate heterogeneity between the set of overall population proportions (*I*^2^ = 64%, *τ* = 0.05). A funnel plot was drawn to assess the publication bias (Figure 4b). The publication bias test results: Egger’s test (p = 0.88) indicated that there was no publication bias.

**Table 3.** All confirmed cases of COVID-19 included in the meta-analysis.

| Study Reference | Age <sup>1</sup><br>(*mean* or<br>median) | Total<br>number of<br>cases <sup>2</sup> | Number of<br>male cases | Number of<br>female<br>cases | All cases<br>(male %) |
| --- | --- | --- | --- | --- | --- |
| Cao et al., 2020 | 54.0 | 102 | 53 | 49 | 52.0 |
| Chen et al., 2020a | 51.0 | 249 | 126 | 123 | 50.6 |
| Chen et al., 2020b | 55.5 | 99 | 67 | 32 | 67.7 |
| Chen et al., 2020c | 54.0 | 203 | 108 | 95 | 53.2 |
| Chu et al., 2020 | 39.0 | 54 | 36 | 18 | 66.7 |
| Du et al., 2020a | 57.6 | 179 | 97 | 82 | 54.2 |
| Easom et al., 2020 | 42.4 | 68 | 32 | 36 | 47.1 |
| Guan et al., 2020 | 47.0 | 1096 | 637 | 459 | 58.1 |
| Huang et al., 2020 | 49.0 | 41 | 30 | 11 | 73.2 |
| Li et al., 2020b | 60.0 | 548 | 279 | 269 | 50.9 |
| Liu et al., 2020 | 57.0 | 137 | 61 | 76 | 44.5 |
| Mao et al., 2020 | 52.7 | 214 | 87 | 127 | 40.7 |
| Qin et al., 2020 | 58.0 | 452 | 235 | 217 | 52.0 |
| Wan et al., 2020 | 47.0 | 135 | 72 | 63 | 53.3 |
| Wang et al., 2020a | 56.0 | 138 | 75 | 63 | 54.3 |
| Wang et al., 2020b | 38.8 | 125 | 71 | 54 | 56.8 |
| Wang et al., 2020c | 50.0 | 1012 | 524 | 488 | 51.8 |
| Wu et al., 2020 | 46.1 | 80 | 39 | 41 | 48.8 |
| Xie et al., 2020 | 60.0 | 79 | 44 | 35 | 55.7 |
| Xu et al., 2020 | 50.0 | 90 | 39 | 51 | 43.3 |
| Yang et al., 2020 | 45.1 | 149 | 81 | 68 | 54.4 |
| Young et al., 2020 | 47.0 | 18 | 9 | 9 | 50.0 |
| Zhang et al., 2020 | 57.0 | 140 | 71 | 69 | 50.7 |

<sup>1</sup> Mean or median age of the study population reported by each research study. Mean ages are indicated in *italics*.
<sup>2</sup> All consecutive patients with lab-confirmed case of COVID-19 within the study period.

### 3.5 Sex-Specific COVID-19 Severe Case Distribution

A total of 8 studies with 985 severe COVID-19 cases were analyzed (Table 4). Our results from the randomized effects model meta-analysis showed that in the sex-distribution of all COVID-19 severe cases, males accounted for 56% (95% CI [0.53, 0.59]) (Figure 5a). Female patients made up 44% of all COVID-19 severe cases. There is no heterogeneity for the severe population proportions (*I*^2^ = 0%, *τ* = 0.0). A funnel plot was drawn to assess the publication bias (Figure 5b). The publication bias test results: Egger’s test (p-value = 0.40) indicated that there was no publication bias.

**Table 4.**
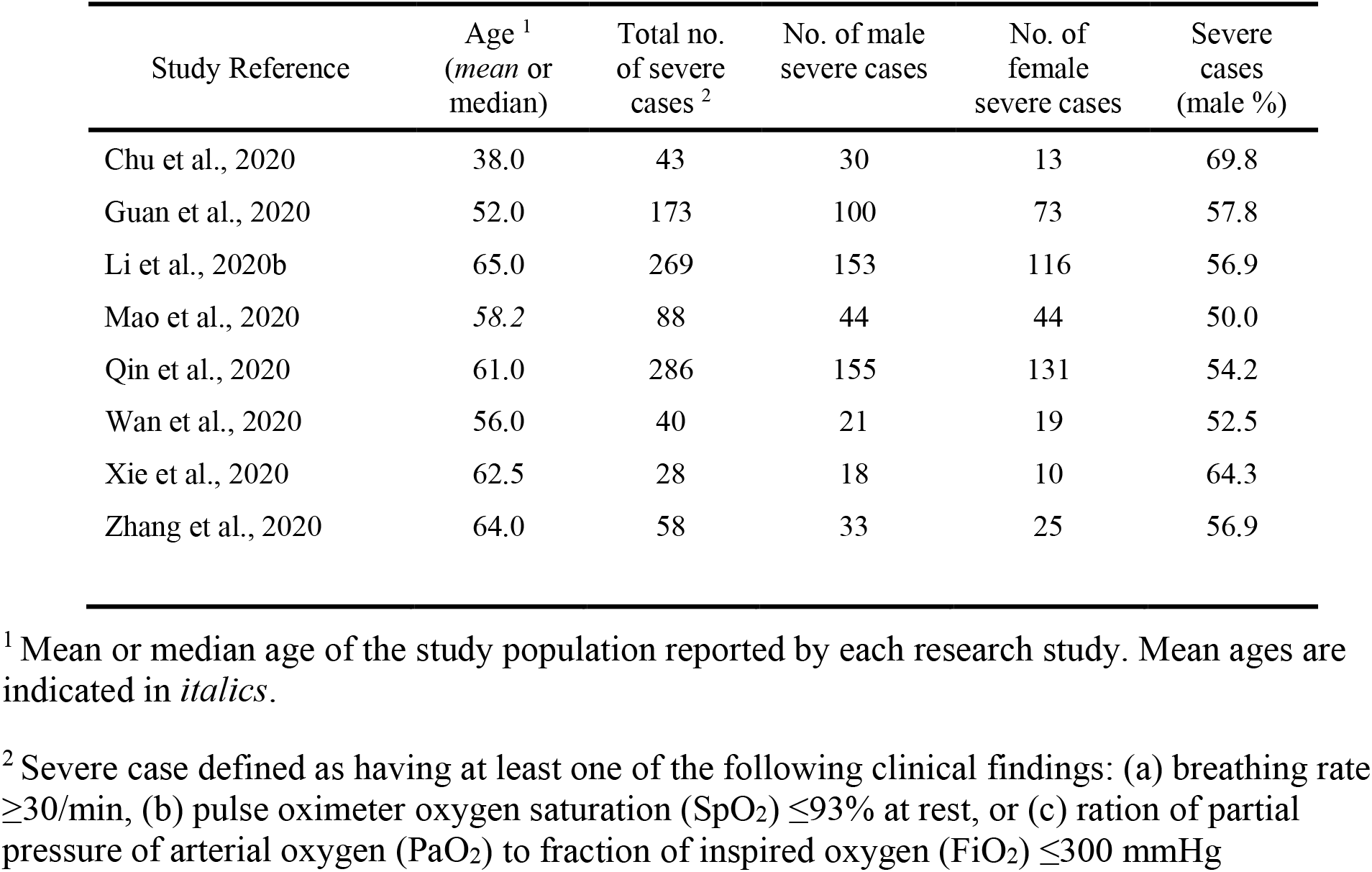
All severe cases of COVID-19 included in the meta-analysis.

### 3.6 Sex-Specific COVID-19 Critically Ill Case Distribution

A total of 9 studies with a total of 2025 critical COVID-19 cases were analyzed (Table 5). Our results from the randomized effects model meta-analysis showed that in the sex-distribution of all COVID-19 critically ill cases, males accounted for 71% (95% CI [0.63, 0.79]) (Figure 5c). Female patients made up 29% of all COVID-19 critical cases. There was strong heterogeneity between the critical population proportions (*I*^2^ = 83%, *τ* = 0.10). A funnel plot was drawn to assess the publication bias (Figure 5d). The publication bias test results: Egger’s test (p-value = 0.02) indicated that there could be some publication bias introduced by the Grasselli et al. (2020) study.

**Table 5.** All critical cases of COVID-19 included in the meta-analysis.

| Study Reference | Age <sup>1</sup><br>(*mean* or<br>median) | Total no.<br>of critical<br>cases <sup>2</sup> | No. of male<br>critical cases | No. of<br>female<br>critical cases | Critical<br>cases<br>(male %) |
| --- | --- | --- | --- | --- | --- |
| Barrasa et al., 2020 | 63.2 | 48 | 27 | 21 | 56.3 |
| Bhatraju et al., 2020 | 64.0 | 24 | 15 | 9 | 62.5 |
| Chen et al., 2020a | 51.0 | 22 | 19 | 3 | 86.4 |
| Grasselli et al., 2020 | 63.0 | 1591 | 1304 | 287 | 82.0 |
| Guan et al., 2020 | 63.0 | 67 | 45 | 22 | 67.2 |
| Huang et al., 2020 | 49.0 | 13 | 11 | 2 | 84.6 |
| Simonnet et al., 2020 | 60 | 124 | 90 | 34 | 75.3 |
| Wang et al., 2020a | 66.0 | 36 | 22 | 14 | 61.1 |
| Wang et al., 2020c | 55.5 | 100 | 62 | 38 | 62.0 |

<sup>1</sup> Mean or median age of the study population reported by each research study. Mean ages are indicated in *italics*.

### 3.7 Sex-Specific COVID-19 Mortality Distribution

A total of 6 studies with a total of 399 mortalities related to COVID-19 cases were analyzed (Table 6). Our results from the randomized effects model meta-analysis showed that in the sex-distribution of all COVID-19 mortalities, males accounted for 69% (95% CI [0.63, 0.75]) (Figure 5e). Female patients made up 31% of all COVID-19 mortalities. The heterogeneity for the mortality population proportions is low (*I*^2^ = 34%, *τ* = 0.04). A funnel plot was drawn to assess the publication bias (Figure 5f). The publication bias test results: Egger’s test (p = 0.26) indicated that there was no observable publication bias.

**Table 6.** All deaths in COVID-19 patients included in the meta-analysis.

| Study Reference | Age <sup>1</sup><br>(*mean* or<br>median) | Total<br>number of<br>deaths <sup>2</sup> | Number of<br>male<br>deaths | Number of<br>female<br>deaths | Mortality<br>(male %) |
| --- | --- | --- | --- | --- | --- |
| Cao et al., 2020 | 72.0 | 17 | 13 | 4 | 76.5 |
| Chen et al., 2020d | 68.0 | 113 | 83 | 30 | 73.5 |
| Du et al., 2020a | 70.2 | 21 | 10 | 11 | 47.6 |
| Du et al., 2020b | 70.7 | 109 | 74 | 35 | 67.9 |
| Du et al., 2020c | 65.8 | 85 | 62 | 23 | 72.9 |
| Korea, 2020 | 75.5 | 54 | 33 | 21 | 61.1 |

<sup>1</sup> Mean or median age of the study population reported by each research study. Mean ages are indicated in *italics*.
<sup>2</sup> All consecutive number of deaths in COVID-19 patients that occurred during the study period.

### 3.8 Sex-Specific COVID-19 Distribution in Asia and the West

Sex-specific differences in clinical outcomes of COVID-19 cases in China were thought to be related to cultural and social differences in males and females (Cai, 2020). We investigated if our study results hold in different regions of the world. COVID-19 critically ill patient data sets were divided into two groups: Asia and West, and subgroup analyses were performed.

A total of 5 studies from Asia, with a total of 238 critical COVID-19 cases were analyzed. Our results from the randomized effects model meta-analysis showed that in the sex-distribution of COVID-19 critically ill cases from Asia, males accounted for 71% (95% CI [0.61, 0.81]) (Figure 6a). Female patients made up 29% of all COVID-19 critical cases in Asia. There was moderate heterogeneity between the critical population proportions (*I*^2^ = 64%, *τ* = 0.0082). A funnel plot was drawn to assess the publication bias in studies from Asia (Figure 6b). The publication bias test results: Egger’s test (p-value = 0.26) indicated that there was no observable publication bias.

**Figure 6.**
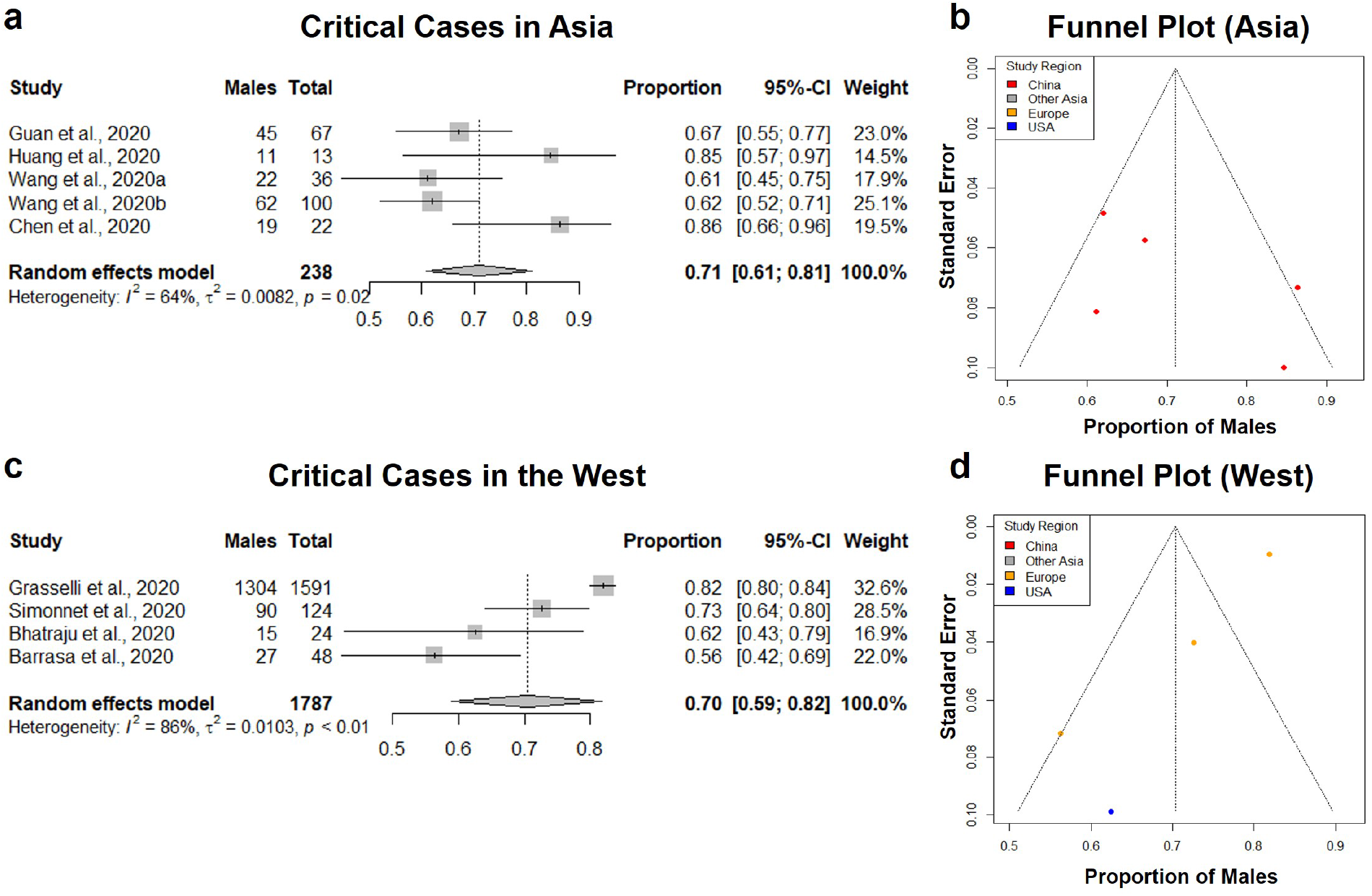
Comparison in the proportion of males in COVID-19 critical cases in Asia and in the West. (a, c) Forest plot of sex-distribution in COVID-19 critical cases in each of the studies. Proportions of males and the 95% confidence intervals are indicated. Critical case defined as: received mechanical ventilation, clinically diagnosed with shock, received care in the intensive care unit (ICU) or transferred to a higher level of medical care. (a) Critical cases in Asian countries. (c) Critical Cases in western countries. (b, d) Funnel plot with 95% confidence region of sex-distribution in COVID-19 critical cases in each of the studies.

A total of 4 studies from Western regions with a total of 1787 critical COVID-19 cases were analyzed. Our results from the randomized effects model meta-analysis showed that in the sex-distribution of COVID-19 critically ill cases from the West, males accounted for 70% (95% CI [0.59, 0.82]) (Figure 6c). Female patients made up 30% of all COVID-19 critical cases in the West. There was strong heterogeneity between the critical population proportions (*I*^2^ = 86%, *τ* = 0.0103). A funnel plot was drawn to assess the publication bias in studies from the West (Figure 6b). The publication bias test results: Egger’s test (p-value = 0.04) indicated that there could be some publication bias introduced by the Grasselli et al. (2020) study, as indicated previously. This comparative subgroup analysis of Asia and the West indicated that there was no geography-specific difference in the proportion of critically ill COVID-19 male patients. However, indicated by the moderate to strong heterogeneity observed, there are likely variations in male proportion between different studies and regions.

### 3.9 Disease severity stratification and age distribution

When extracting male and female proportions for each of the four COVID-19 disease severity categories, we obtained the age distributions of the cases stated as a mean ± SD or median and interquartile range (IQR). Using a skewed distribution assumption, the ages were aggregated as medians with 95% confidence intervals. The median age for all COVID-19 cases was 50, severe cases was 61, critically ill cases was 63, and mortality was 70 (Figure 7). A Kruskall-Wallis ranked-sum test conducted on the medians showed that age was significantly different between the COVID-19 disease severity groups (chi-squared = 24.07, df = 3, p-value = < 0.0001). Our data confirm that advanced age is a risk factor for more severe clinical outcomes and mortality related to COVID-19.

**Figure 7.**
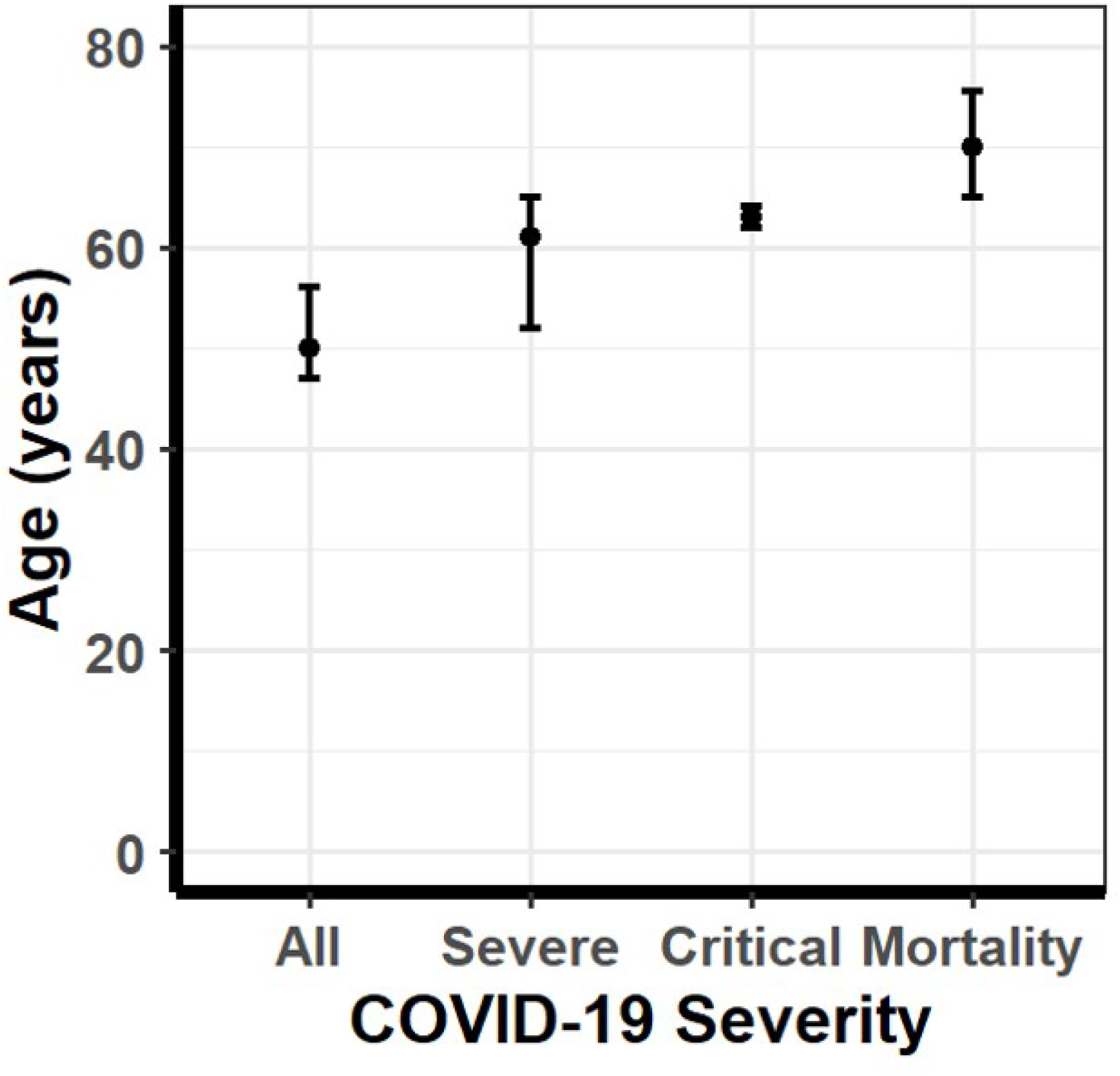
Median age of COVID-19 patients stratified according to disease severity. Median age of COVID-19 patients in all cases, severe cases, critically ill cases, and mortalities. Error bars represent 95% confidence intervals of the median. The median age for all COVID-19 cases was 50, severe cases was 61, critically ill cases was 63, and mortality was 70. A Kruskall-Wallis ranked-sum test conducted on the medians showed that age was significantly different amount the COVID-19 disease severity groups (chi-squared = 24.07, df = 3, p-value = <0.0001).

## 4. Discussion

In our systematic review and meta-analysis, we set forth to address the question of whether male sex is a risk factor for COVID-19 susceptibility, severe forms of the disease, or mortality related to COVID-19. Systematically reviewing all literature from December 15th, 2019, to April 16th, 2020, we selected 31 research studies that met our selection criteria and performed a meta-analysis on COVID-19 clinical outcomes. Our quality assessment measures indicated low heterogeneity in terms of a single-arm meta-analysis, and the sensitivity analysis showed that there was minimal publication bias. As of the time of completing this manuscript, there were no randomized controlled trials with COVID-19 patients that could address this particular question. The use of non-randomized studies for the meta-analysis is a limitation of this study. However, Abraham et al. (2010) suggested that, in the absence of randomized, controlled trials, that a well-designed meta-analysis is used non-randomized controlled trials can also present a high level of evidence (Abraham et al., 2010).

The four clinical outcome categories (overall, severe, critical, mortality) exhibited different levels of heterogeneity in our random-effects models. The explanation for these differences is most likely the region of the studies done within each category. The 23 overall studies exhibited 64% heterogeneity with one from Singapore and one from Great Britain. The eight severe studies exhibited 0% heterogeneity, all being from China. The nine critical studies exhibited 83% heterogeneity, with five from China, three from Europe, and one from the United States. The six mortality studies exhibited 35% heterogeneity, with five from China and one from Korea. The use of a randomized effects model for our meta-analysis takes into account these heterogeneities observed between different studies and regions. Based on the random-effects models shown, there appears to be a difference in the proportions of males with COVID-19 between at least some of the studies or regions. Due to the study designs, their sampling methods, and limited regions included in this study, it is neither possible nor wise to be more specific. This is a potential avenue for further research.

Our meta-analysis showed that while males accounted for 53% of all COVID-19 cases, males accounted for an increasing proportion of severe cases (56%), critically ill cases (71%), and mortalities (69%) compared to their counterpart. While similar male to female disproportions was observed among a few other studies looking at clinical characteristics of COVID-19, our study provides a comprehensive synthesis of data available across different world regions. This study helps establish male sex as a risk factor for COVID-19 clinical outcomes and shows that it is consistent in Asia and Western regions.

This study results do not come as a surprise. Several studies conducted on the two previous coronavirus epidemics, SARS CoV-1in 2002-2003 and MERS in 2012-2013, showed similar patterns with a male predominance toward greater severity and mortality risks. Studies on mortality rates during the MERS-CoV epidemic showed the male sex to be a risk factor (Matsuyama et al., 2016; Nam et al., 2017; Park et al., 2018). Epidemiological studies with SARS-CoV-1 showed similar patterns (Karlberg et al., 2004). To further support previous epidemiological observations, in controlled mouse model experiments, SARS-CoV-1 has displayed infectious dose-dependent higher mortality rates in male mice compared to female mice (Channappanavar et al., 2017). The mounting amount of evidence showing differences among males and female clinical outcomes to coronavirus infections highlights the importance of patient sex in determining the COVID-19 prognosis.

From a clinical standpoint, this information is very pertinent to the practice of patient care. As COVID-19 clinical outcomes are strongly associated with male sex, this can help guide preventative and treatment strategies. Male patients will likely warrant more aggressive inpatient care measures, and especially those that have other COVID-19 risk factors such as advanced age or underlying comorbidities. Susceptible males with other known risk factors may need to take extra precautions to help prevent SARS-CoV-2 infection. Infected males can be encouraged to obtain medical care at an earlier stage of the disease. In cases that require hospitalization, physicians can be more aggressive in their medical management of male patients.

In addition to preventative and COVID-19 treatment measures, this presents a unique clinical opportunity to address male and female differences at the molecular level, immunological response, and endocrine function (Sandberg and Ji, 2003; vom Steeg and Klein, 2016; Taneja, 2018). For example, SARS-CoV-2 binds to the Angiotensin-converting enzyme 2 (ACE2) receptors and use it as a mechanism for host cell entry (Hoffmann et al., 2020). Males have been shown to express more ACE2 receptors within the renin-angiotensin-aldosterone system (RAAS) (Komukai et al., 2010). This is likely to play an essential role in the severity of this disease observed in males (Komukai et al., 2010). Differences in male and female immunological responses will also be a clinically significant factor that can be appropriately modulated to better serve COVID-19 patients (Pennell et al., 2012; Jaillon et al., 2019). Besides sex-specific differences in immunological responses, hormonal regulation and the role of estrogen and testosterone in priming the ACE2 receptor sensitivity could hold the key to better explain the higher COVID-19 severity and mortality rates observed in males (Mishra et al., 2016; Bukowska et al., 2017). In an age of personalized medicine, if molecular level of differences in the disease processes of SARS-CoV-2 infection can be characterized, clinicians can use targeted therapy using to promote health equality and help save more patient lives.

## Data Availability

All datasets presented in this study are included in the article or supplementary files.

## Acknowledgments

We acknowledge Dr. Siva Somasundaram, Ms. Avishka Jayasekera, and Dr. John P. Walsh of the University of Southern California for their collaborative support. We also thank Dr. Jeffrey S. Wang, Infection Disease Specialist at Kaiser Permanente, Anaheim, California, for his clinical insights.

## Author Contributions

TG led the systematic review, helped prepare the tables and figure, and aided in writing and editing. BMP aided in the analytical evaluation of curated articles, writing, and editing. JA, AB, and DR research students analyzed data and joined in discussions. JW performed statistical analyses; drafted statistical sections. RSG conceptualized the problem and aided in writing and editing.

## Supplementary Material

Supplementary Materials are provided as a separate file.

